# Translating motivations, barriers, support and information into health behaviour change: community members’ response to Australia’s National Preventive Health Strategy

**DOI:** 10.64898/2026.08.12.26358631

**Authors:** Amie Steel, Hope Foley, Jon Adams

**Author notes:** Corresponding author: Dr Hope Foley, NCNM, Southern Cross University.

## Abstract

Preventive health is a crucial health systems component for managing disease burden and achieving health promotion policy goals. However, effective prevention relies on the modification of relevant risks, often requiring systemic health behaviour change. Australia’s National Preventive Health Strategy (NPHS) prioritises seven focus areas: tobacco and nicotine, healthy diet, physical activity, cancer screening, immunisation, alcohol and other drugs, and mental health. The readiness of community members in Australia to address health behaviours relating to these areas has not been fully examined. In response, six focus groups were conducted with 27 adults from the Australian general population to explore their perspectives and experiences of preventive health information and behaviours relating to the seven NPHS focus areas. Themes and sub-themes were identified using an applied descriptive framework. Participants described motivations, barriers and experiences surrounding preventive health through the themes of ‘Making informed health choices’, ‘Facilitating behaviour change and the role of support systems’ and ‘Spreading the preventive health word’. Sub-themes detailed processes of prioritisation, risk-benefit assessment, critical appraisal, sociocultural influence and support-seeking to understand and personalise preventive health information, implement behavioural change, and share information with others. The focus areas participants engaged with most strongly were healthy eating and physical activity, while cancer screening was discussed less often. These findings indicate high preventive health engagement in the Australian community, alongside challenges navigating and adapting relevant information to personal needs. These insights can support policymakers, healthcare providers and others to effectively enact the NPHS through more targeted preventive health information and care delivery.

## Background

Preventive health is recognised as one vital element of contemporary healthcare [1]. A preventive health approach addressing health risks before they affect health is relevant to all health conditions including non-communicable diseases, communicable illness and even adverse events associated with the management of diagnosed illness [2]. While prevention is known to be a highly cost-effective method of improving population health in the short, medium and long-term [3], its success relies heavily on behavioural factors and, subsequently, on facilitating and sustaining population-level changes in health behaviour.

Health behaviour change relies on complex, intersecting internal and external factors. For example, an individual’s health behaviours are first defined by available options based on structural (e.g., service provision and costs), socio-cultural (e.g., housing, language), economic (e.g., education, income), commercial (e.g., marketing, supply chains) and environmental (e.g., climate change, green spaces) determinants [4]. These determinants largely affect individual capability to change behaviour [5], while the options they choose are further influenced by their beliefs, which in turn drive motivation and intention [6]. The Australian government’s first National Preventive Health Strategy (NPHS) acknowledges these wide-ranging factors as critical to preventive health success [4]. The NPHS also outlines seven focus areas to prioritise: tobacco and nicotine, healthy diet, physical activity, cancer screening, immunization, alcohol and other drugs, and mental health. The intersection between these external and internal factors and the role they play in the population’s engagement with the breadth of behaviours targeted in the NPHS focus areas, however, is not clearly understood.

Beliefs are significant internal factors influencing health behaviours due to their effect on motivation and intention [7]. One of the primary precursors to belief is knowledge, and consequently, access to accurate, timely and relevant health information is critical to supporting effective health behaviour change. The contemporary health information landscape is complex and this includes (but is not limited to) preventive health topics such as diet [8], immunisation [9] and alcohol use [10], with recent research suggesting the Australian population may have knowledge gaps regarding some of these crucial preventive health topics [11]. The community’s expressed preventive health information needs emphasise the imperative role of health promotion agencies and practitioners in disseminating appropriate information. However, to reach the community and achieve preventive health objectives such messaging must compete with the diverse messages and information sources currently dominating the health information landscape [12, 13]. Beyond informing the community, health promotion principles also recognise the importance of supporting individuals to overcome external barriers (e.g., structural, social or economic factors) to implement behaviour change [14]. As such, successful implementation of the NPHS requires closer examination of the community’s perceptions and experiences of learning about and implementing change to address Australia’s preventive health priority behaviours.

## Methods

### Study design

#### Study Aim and Framework

This study employed in-person and online focus groups, and an applied descriptive framework [15] to explore the perceptions and experiences of the general population towards accessing and using information about the preventive health focus areas of the NPHS. The study was conducted in accordance with the Helsinki Declaration, and ethics approval was granted by the University of Technology Sydney Human Research Ethics Committee (ETH24-9111).

#### Participant selection

Participants were recruited between 24 April and 29 July, 2024, using purposive recruitment via social media advertisements on Meta platforms (Facebook, Instagram) and X/Twitter. Most advertisements targeted adults (aged 18 years and over) in Australia focusing upon specific locations where in-person focus groups were planned (e.g., Sydney, Brisbane, Hobart, Melbourne). Other advertisements targeted adults beyond those locations for participation in online focus groups. A brief online consent and expression-of-interest form identified basic demographic and healthcare use information to allow purposive recruitment across various characteristics. A total of 27 participants contributed to the focus groups. The characteristics of participants are described in Table 1, while further details of recruitment and sampling are available in the supplementary information file (see Supplementary Figure S1 and Supplementary Table S1).

**Table 1.** Demographic characteristics of participants and focus group locations.

|  | N | % |
| --- | --- | --- |
| **Whole sample** | 27 | 100 |
| **Gender** |  |  |
| Female | 14 | 51.9 |
| Male | 11 | 40.7 |
| Non-binary | 2 | 7.4 |
| **Age** |  |  |
| 18-25 years | 4 | 14.8 |
| 26-35 years | 9 | 33.3 |
| 36-45 years | 5 | 18.5 |
| 46-55 years | 3 | 11.1 |
| 56-65 years | 4 | 14.8 |
| 66-75 years | 2 | 7.4 |
| **Focus group location** |  |  |
| Melbourne | 8 | 29.6 |
| Sydney | 6 | 22.2 |
| Brisbane | 4 | 14.8 |
| Hobart | 4 | 14.8 |
| Online | 5 | 18.5 |
| **Participant state** |  |  |
| VIC | 10 | 37.0 |
| NSW | 6 | 22.2 |
| QLD | 6 | 22.2 |
| TAS | 4 | 14.8 |
| WA | 1 | 3.7 |
| **Participant locality** |  |  |
| Capital city | 23 | 85.2 |
| Regional or rural | 4 | 14.8 |

#### Setting

In-person focus groups were conducted in private rooms in community locations such as libraries and community centres, except the Sydney focus group which was conducted on campus at the University of Technology Sydney (Ultimo, Sydney). Online focus groups were conducted using Zoom.

#### Data collection

The focus groups followed a semi-structured format allowing a schedule of core discussion topics to be covered while remaining responsive to new topics introduced by participants in the field. An infographic of the seven priority areas from the NPHS was provided to participants as a prompt when discussing preventive health topics. The discussion guide and infographic are available in the supplementary information file (see Supplementary Methods S1). All discussions were audio recorded with participants’ consent and supplemented by note-taking. The focus groups ran for between 55 and 98 minutes with an average of 74 minutes. In accordance with data saturation, we completed a total of six focus groups (four in person, two online).

#### Data analysis

Data were initially coded by AS with input from HF and JA using the Framework Method [16]. Themes were documented using NVivo 14 [17]. All three facilitators revisited the coding independently and together to verify alignment with the focus group discussions.

## Results

Three major themes emerged from the analysis of the focus group data: (1) *Making Informed Health Choices*; (2) *Facilitating Behaviour Change and the Role of Support Systems*; and (3) *Spreading ‘Preventive Health’*. These themes and their subthemes are presented in Figure 1.

**Fig. 1.**
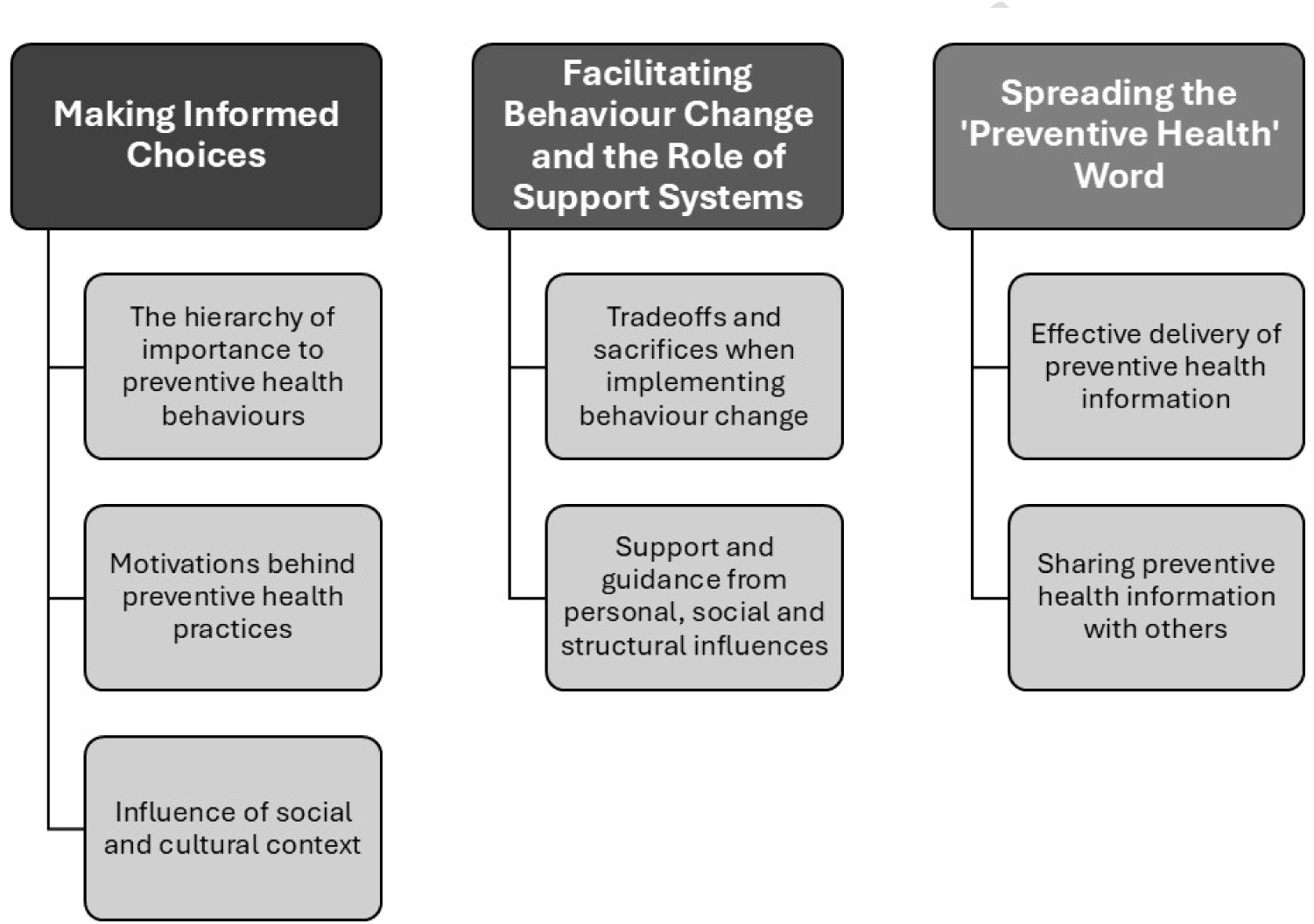
Themes and subthemes identified during applied descriptive framework analysis

### Making informed health choices

Participants described their engagement with and adoption of a range of preventive health behaviours and practices, including those which the categories outlined in the NPHS. Participants were keen to place differing degrees of importance on different health behaviours and described the motivations behind these behaviours.

#### A hierarchy of importance of preventive health behaviours

Participants were encouraged to discuss their engagement with health practices to achieve or maintain health. All participants reported some preventive health practices, placing varying degrees of emphasis on different behaviours. Physical activity and healthy eating dominated many accounts with mental health, alcohol, smoking, and immunisation also discussed. Cancer screening was not raised as an important area of preventive health.

Participants across all focus groups identified physical activity as part of their personal health regime. However, the degree and form in which they engaged varied. Some participants described establishing specific detailed exercise programs or activities separate and distinct to day-to-day activities. Others outlined a ‘low level’ integration of physical activity alongside longstanding habits, for example adapting everyday travel routines to incorporate physical activity where possible. A few participants explained combinations of these approaches.

Healthy eating was another preventive health topic raised by many participants across all focus groups, with specific issues outlined as indicators of better food choices and consumption. Many participants perceived healthy eating as synonymous to home-prepared food rather than ready-made store-bought or takeaway meals. Linked to the importance of home-cooked food was that of meal planning, although this was not always considered a recipe for successful healthy eating. Other participants emphasised that portion size affected whether they were eating healthily even when cooking at home. Typically, participants viewed healthy eating behaviours in general terms and were somewhat dismissive or sceptical of specific diets. Alongside physical activity and healthy eating, participants also explained how engaging in preventive health practices – some of which were explicitly linked to physical activity (e.g. bushwalking, yoga) – help to also boost their mental health.

Less common, but still mentioned by participants as preventive health practices, were avoiding alcohol, tobacco smoking and other substances. Total abstinence from smoking was reported as the ideal health behaviour by all participants who discussed smoking, either as something they used to do but have stopped or something they have never done, and in one instance was grouped with other ‘drugs’. Alcohol, meanwhile, was commonly described as something ideally avoided but often still consumed, with the aim to limit excess.

#### Motivations behind preventive health practices

In addition to outlining their chosen preventive health practices, participants often provided reasons for engaging in those practices. These motivations mostly included *managing specific health concerns*, but also related to *ageing and longevity*, *general wellbeing and quality of life*, and *responding to social and cultural norms*.

##### Managing their personal risk of specific health conditions

Participants in each of the six focus groups described the importance of preventive health practices to manage a specific health condition. In some instances, such as an older female participant concerned about falls risk, they are motivated to prevent a yet to be experienced specific health condition they feel poses a significant personal risk. This particular participant describes how she uses physical activity to manage her risk of falls and prevent related health problems:

> “*I’m quite aware of things such as falls…If you lose your muscle, you become weak, you fall. If you fall once you’re likely to fall again. There’s that vicious cycle. Once you have a fall then depending on whether you have an injury, you might be confined to bed for a long time*.” – Melbourne, Female, 56-65 years

Another participant described being similarly motivated to adopt healthy eating to help prevent diabetes, which she identified as a high personal risk:

> “*I really try to eat healthy. So, I had a near diabetes scare…I have polycystic ovarian syndrome, which is not diabetes, but it puts me at a high risk. So, that was enough. So, I was quite overweight, not quite obese, but very near the line. I’m like, ‘If I don’t change my lifestyle now, I will most likely get diabetes by the time I’m 30.’*” – Online FG2, Female, 26-35 years

A third participant expressed a similar view around preventing future health problems through implementing diet changes in response to concerns, raised by his doctor, about his weight:

> “*Well, I guess a few months back I went to the doctors and did a whole health checkup, and he didn’t say anything was wrong, but he just said, ‘You need to stay healthy and to reduce your weight, because it’s like an endemic. And people eat all sort of sugary foods, so you need to reduce your weight.’ And yeah,…since then I’ve actually reduced five kilos*.” – Hobart, Male, 26-35 years

Highlighting the impact of this sense of personal risk regarding a specific health condition was a participant who described only being motivated to access some types of cancer screening if they had a personal experience with the cancer, either through their own health or someone they know:

> “*The only reason I would get screened for certain cancer types is because I know someone or I’ve had a personal experience with that*.” – Hobart, Female, 26-35 years

Another participant described how they would take an approach more in line with secondary health prevention, whereby they would only act if experiencing symptoms requiring attention:

> “*I work on the basis of if I’m not experiencing health symptoms or problems, if this isn’t a problem, then whatever I’m doing couldn’t be that bad*.” – Brisbane, Non-binary, 26-35 years

##### Addressing ageing, longevity, general wellbeing and quality of life

Participants also identified the more general issues of longevity, wellbeing and quality of life as motivations for engaging in preventive health behaviours. While prevention of risks associated with ageing were mostly raised by older adults (60 years and older), one younger participant in their late 20s also identified concerns about ageing as motivating their decision to engage in preventive health behaviours:

> “*I got to [my late 20s] and I’m like, I don’t want get to 40 and look 50. I want to get to 40 and look 30…I’ve had my wild years in my early twenties and now it’s like ‘start to think more long term’..*. *So yeah, that was probably the main thing was just the age and stage of life. That was just the point where I started thinking … wanting to go into my 30s with better lifestyle habits.*” – Brisbane, Non-binary, 26-35 years

Other participants described attempting certain health practices such as healthy eating, physical activity and reduced alcohol intake merely because they knew they were healthy behaviours or had noticed small ways they made them feel healthier and improved their quality of life overall. One participant, for example, explained that simply knowing a behaviour had health benefits was enough motivation for them to try it:

> “*I would say I haven’t forced myself to do it, but it’s just something I know that it’s good for my health anyway, so yeah, why not? So just try it*.” – Brisbane, Male, 36-45 years

Other participants gave direct examples of how unhealthy behaviours affected their cognitive function at work or their physical performance in their hobbies and how this motivates them to choose healthier behaviours:

> “*Once you’re … just doing stuff three to four times a week you might think like, hey, if I’m drinking and smoking all the time, I’m probably not going to be as smart. If I’m going out drinking until 3:00 AM in a lot of nights in a row when I have important work, it’s just going to impact what you want to do*.” – Brisbane, Male, 18-25 years
>
> “*Just realizing the impact it would have to drink and then try and do things the next day, and…that impact it has on my health and being able to train…, I’ve just realized it’s not for me*.” – Hobart, Female, 36-45 years

#### Influence of social and cultural context

Participants acknowledged the influence of the wider social and cultural context upon their preventive health practices, primarily in relation to their living situation. One participant, for example, described how his physical activity level was influenced by a current romantic relationship, indicating they will put effort into ‘getting fit’ when their girlfriend is overseas:

> “*I’m in my young twenties, got a girlfriend at the moment, but she’s probably going back to Japan in the next month, I think I’ve probably got to get fit again*” – Brisbane, Male, 18-25 years

Another participant suggested his wife’s veganism influenced his own diet.:

> “*I don’t get any meat or chicken. Or get very little meat or chicken unless we go to a restaurant. So, I eat a lot of lentils, and once again, but it’s good. I feel really good for it.*” – Sydney, Male, 66-75 years

Other participants described broader socio-cultural influences - such as religion, as was the case for one participant, and family alcohol abuse in the case of another - as informing their decision not to drink alcohol:

> “*For me, I mean it’s a cultural and religious aspect, so we don’t drink alcohol so it’s out of the question*” – Hobart, Male, 26-35 years
>
> “*Probably seeing through the impact of alcohol on family and things like that is also probably a big piece [of personal alcohol avoidance] as well*.” – Hobart, Female, 36-45 years

Another participant also social influences in terms of responding to a perceived general social movement towards healthy behaviours. Meanwhile, other participants mentioned being influenced by wider determinants such as concerns about the environmental impact of their diet:

> “*My healthy obsession started probably seven years ago with the decision for my environmental principles to become vegan in my diet*.” – Online FG1, Male, 46-55 years

### Facilitating Behaviour Change and the role of Support Systems

Participants shared their views on the factors that facilitate their own behaviour change. These included *tradeoffs and sacrifices when implementing behaviour change*, and *the role of personal and structural support systems*.

#### Tradeoffs and sacrifices when implementing behaviour change

Participants identified factors that influenced their decision, or ability, to change their health behaviours, requiring what they perceived as trading off benefits against sacrifices.

##### Ease of implementation

Several participants described the ease of implementing behaviour change as critical to making a positive change:

> *“For me, when you look into more information, I mean what I do is I think it’s human nature. I try to find what’s easiest for me to adopt and then I go for that.”* – Hobart, Male, 26-35 years

Similarly, another participant highlighted the challenge of cold weather upon their healthy eating behaviours:

> *“I find it really difficult as the weather starts to get colder to be better prepared at home. Because once you get home you don’t want to go out again and it’s like, ‘Okay if I don’t have everything here.’ So yeah, I’ve certainly been making more of an effort to make sure I’ve got a fridge full of stuff rather than just a handful of things.”* – Hobart, Female, 56-65 years

Other participants explained that maintaining a healthy diet (through effective preparation) could come with unavoidable trade-offs, whether this was limiting food choices or compromising on taste:

> *“The other thing that I do is I try and minimise the friction in making the food. If I’m cooking, I’ll cook for four straight meals, although I understand that I’ll have to eat the same thing for four straight meals, but I don’t mind, really, because that’s a trade-off I’m willing to make in order to eat healthy.” -* Melbourne, Male, 26-35 years

##### Cost savings and expenses of healthy behaviours

Some participants recognised financial circumstance as a challenge requiring a trade-off on their health behaviours and affecting their ability to undertake (amongst other things) healthy eating, cancer screening and mental health management.

> *“If people are on Centrelink, they can’t necessarily afford the healthier food options.”* – Brisbane, Non-binary, 26-35 years
>
> *“So for me, the skin cancer, I want to go and get that checked, but it costs me money to get it. And so every time I deprioritize it.” –* Hobart, female, 56-65 years
>
> *“Everyone tells you can get 10 free sessions to see it, but there’s no psychologist that is gap free at all here. So, if anyone wants those services, if you can’t pay for them, you’re not getting them.*” – Hobart, Female, 56-65 years

However, in the case of healthy eating, one participant also described how meal planning and preparation helped them save money through avoiding junk food:

> *“I’m a student. What I do is I usually have my next two meals planned. I have it cooked in my refrigerator so that I know that I won’t be eating junk. The other thing that I do is before going to uni, I make it a point to have something packed. It obviously helps me cut down on expenses, but that also ensures that I don’t eat anything junk.”* – Melbourne, Male, 26-35 years

A second participant explained the money they saved by eating at home allowed them to spend more on organic food, which they perceived as providing health benefits:

> *“I prioritize organic food…I only eat organic food except if I go to a restaurant. And so that’s a price, it’s a high price, but also, I’m cooking…95% of my meals at home are from scratch. So hopefully, saving money on pre-prepared foods.”* – Online FG1 – Male, 46-55 years

This same participant also mentioned the importance of finding ‘low cost’ ways of engaging in physical activity – such as cycling or running rather than going to the gym:

> *“I enjoy not going to the gym, but I have some weights at home and a yoga mat and I go running, and … I cycle. And basically, the low cost, I do love the fact that running is just a pair of shoes every now and then. And the yoga mat’s lasted me for about maybe 14 years now, and maybe will keep going, whatever.”* – Online FG 1, Male, 46-55 years

##### Enjoyment of life is important too

For some participants, life enjoyment was an element they considered when deciding which behaviour change to implement, and the degree to which they should implement it. As two participants in the Brisbane focus group explain:

> “*P1: At the end of the day, we don’t live with the sole purpose and intention of being as healthy as possible. Generally, we think if someone did that, it’s a depressing way of life.. You want a balance of these things. You want to have enjoyment as well*.
>
> *P2: I think it’s like in moderation. You can consume alcohol, you can eat some junk food, for example, if it’s not too much. As [P1] said, this is for enjoyment in life as well*.” – Brisbane, Non-binary 26-35 years [P1] and Male 36-45 years [P2].

A related perspective was shared by a participant in Sydney who indicated healthy behaviours were important as a way of making her happy, so she avoided healthy foods that did not support this happiness (even if recommended by health professionals):

> “*So for me, if some professionals give me some advice, give me a list what you should eat or not to eat…I have a look because I’m eating some healthy stuff to make me happy. If they want me to eat something I don’t want, I will not take it.”* – Sydney, Female 46-55 years

#### Support and guidance from personal, social and structural influences

Participants outlined several personal, social and structural factors they felt provided the necessary support and guidance to successfully implement preventive health behaviour changes.

The most common support identified was that provided by health professionals including specialist medical doctors, general practitioners, dieticians and nutritionists, psychologists, osteopaths, chiropractors, naturopaths, physiotherapists and personal trainers. In general, health professionals were seen as a useful resource to be approached judiciously, when expert knowledge is considered appropriate or essential:

> *“I think it depends on the topic. If it’s something general like eat healthy, eat more fruit, eat more vegetable, that’s something easy I can adapt to my lifestyle. But something very specific like, let’s say, to take this medicine but need to make sure it doesn’t have a effect with the other vitamin that I’m taking. So something that I’m not sure right now whether I should follow this instruction or should I consult the doctor.”* – Brisbane, Male, 36-45 years
>
> *“So, personally I don’t go to the doctor as much because I think generally I’m healthy and only if something really bad has happened then I would want to go.” –* Melbourne, Male, 18-25 years

##### The ideal versus the practicalities of accessing health professionals for preventive health

Participants described different reasons for accessing each of the diverse range of health professionals for support with health behaviour change. While general practitioners were a major focus for seeking professional preventive health advice, many participants also expressed frustrations and challenges in accessing and receiving helpful information from their general practitioner, as the following quotes highlight. Respectively, these participants below note particular challenges associated with: limited time to talk with the doctor in short consultations; certain restrictions in appointment booking systems; doubting the ability of the doctor to provide useful preventive health advice due to the doctor’s apparent conservative perspective; and having a previous negative experience with their doctor.

> *“I tend to find there’s never enough time with the GP, no matter what. I’m writing things down so I don’t forget when I see them. Because you’ve got this such a short period of time and because I don’t go very often …so, try and get everything in together. … I find a lot of things kind of just get [forgotten] about because it just feels like there’s not enough time and there’s not enough resources to see them again.”* – Hobart, Female, 56-65 years
>
> *“In the last 10 years I actually moved to another suburb. I found that some GPs in my new suburb, they may be quite busy so they even would make a sign, that if you make a standard appointment you can only ask one problem. If you have two problems, you have to book like a longer appointment, things like that. So, it just makes me feel like I don’t think it’s appropriate to even ask other things or just ask for recommendation,*.” – Melbourne, Female, 46-55 years
>
> *“There are very few doctors that are progressive and that’s the problem. And I think that’s where we’re told this is the way you prevent this particular health condition and this is the way you do this, and the traditional sort of ways of doing it” -* Online FG1, Male, 56-65 years
>
> *“I had a really bad experience with my GP recently and that’s really put me off…So now I’ll just pay and go and see an [osteopath] off my own back without the referral because it’s easier. So, I’ve kind of cut my GP.”* – Hobart, Female, 36-45 years

##### Preventive health information needs to be tailored before it can be implemented

Some participants described the importance of information from health professionals being personalised - a tailoring that participants received from different health professional groups to varying affect. One participant explained health professionals often provide very general information on preventive health which leaves the patient to personalise that information to enable action on it:

> *“I think the issue is, personally for me, I think it was a point where I realized that GPs and all these health professionals can give me that certain advice, but it’s not personalised to me. They give very general advice because of the time constraints. So, it is up to me to personalise it. So, I think, for me, it’s a lot about not just them giving me information, but teaching me how to get to that point of, now, I know.”* - Melbourne, Male, 18-25 years

In response, participants discussed accessing care they considered more specialised from allied health professionals who were able to personalise advice, such as this participant who preferred dietician advice to guide their healthy eating:

> *“In my experience I feel like GPs gave me very incomplete or very unpersonalised things. It’s like look ‘that’s not sustainable because for me I used to be an athlete’. It’s like ‘what you just told me is not what athletically sustainable for me.’ I need a high carb diet to do my squats, things like that. So, I would trust a dietician.”* – Melbourne, Male, 18-25 years

While other participants described accessing a range of non-biomedical health professionals for preventive health care:

> *Where [other participant] was talking about the Chinese medicine and I might do reflexology or I might do massage or I might do meditation. And those things are great for dealing with pain and also for preventative. So I think there’s got to be a mix.”* Online FG1, Male, 56-65 years

Others described enlisting other practitioners and informal networks to help tailor any advice received from professional sources:

> *“There are some people I would trust as well that I know in my own life, like I’ve got a mate who’s a nutritionist. So sometimes I’d ask him like, “Hey, is this actually a healthy meal plan?” Or same thing, I got mates who are personal trainers. So, if I was going to do something in the gym, “Hey, what’s a good routine?“* – Brisbane, Male, 18-25 years.

##### Government has a role in facilitating access to preventive health care

Participants often described the government as responsible for providing support or guidance on community preventive health behaviour change. Most notably, government was seen as responsible for funding access to services which assist participants to receive the personalised, expert advice they felt they needed to implement preventive health behaviours. This participant from Hobart, for example, described feeling it was disingenuous and economically problematic for the government to advocate for individuals to engage in preventive health behaviours without funding support:

> *“If [the government] are going to promote it, I feel like it needs to be also backed in to go ‘and how are you going to support us to have access to that?’ It’s very shortsighted economically not to invest in those things because a healthy country means healthy economy …my feeling is if you can’t back it into support how you’re actually going to implement it, then I’m skeptical of what they’re saying if it’s, if they can’t support it.”* – Hobart, Female, 36-45 years

This point was furthered by a fellow participant who described finding information online but then requiring additional support to use the information:

> *“But when you visit those websites, they provide you simple things to do. But actually if you are really in need of help, you need to actually meet someone and you just don’t need words out of them, you need some help… But if for the government, because they’re providing healthcare, so if they could provide just to take the next step in keeping the public healthy or promoting a natural lifestyle, free nicotine patches for the first few months, just to get that habit of… Instead of just saying it, what you need to change, you actually are part of change.”* – Hobart, Male, 26-35 years

Other participants specifically suggested GPs be funded to meet their obligation to discuss preventive health topics such as alcohol and drug use with their patients:

> *“At the moment, GPs are provided with incentives for things such as immunization and I do believe that for preventive health things such as alcohol, drugs, I think… And given that some of the info sources can be a bit difficult for some people to understand, digest, I think it would be very helpful for GP… In fact, it should be an obligation for certain health issues for them to talk about preventive steps because we are already doing it with immunization.”* – Melbourne, Female, 56-65 years

And another participant:

> *“I think it would be really good if the governmental authorities, they could …compile a certain amount of information, where they think we can trust. And so when we have a little bit of questions, we can go for those information as an initial consultation. Then, we can bring those information, all of the questions that we have, to follow up with the healthcare practitioners.”* – Melbourne, Female, 26-35 years

### Spreading the ‘preventive health’ word

Preventive health information was deemed by participants as having to be presented in a certain way for it to effectively reach them, and for them to feel able to share the information with others.

#### Effective delivery of preventive health information

According to participants, the preventive health information most effective at reaching community and instigating behaviour change is relatable, repeated, opportunistic, visual, fun and legitimate.

##### Peer advice is more relatable and relevant

Several participants provided examples of valuing preventive health information sourced from a peer, which they often found especially relevant to their own life. Such advice could be from a friend or colleague, or from a more formal peer. Either way it was important the person providing the advice had some experience (and success) applying the information in their own life and could share their personal experiences, as described by this participant who found peer support through Narcotics Anonymous particularly useful in addressing substance use:

> *“You definitely go to somebody who’s been in your shoes, who understands what you’ve been through and who has gotten themselves out of it. If you go to a medical professional they’re going to be like, ‘Get off drugs, what are you doing on drugs?’ Well, obviously I don’t want to be. Thank you for that great advice. If I could just go, okay, no, thank you. None today. No, it’s not that simple. You got to go to the people that have been in your situation before you and have gotten through it.”* – Brisbane, Female, 26-35 years

##### Legitimate sources mean more

Participants also perceived value in receiving the same information repeatedly, particularly if provided in different settings, social media platforms or from organisational accounts. However, as described by this participant, seeing something from an organisation they deemed trustworthy would influence them to pay fuller attention even if it had been repeated elsewhere:

> *“If I had a Queensland Health or an ABC [public broadcaster] thing come up, it’s more like you scroll a bunch of stuff you don’t remember, but if something pops out you go, oh really? Then I’ll look more into it.” –* Brisbane, Male, 18-25 years

##### ‘I need to really visualise it’

Some participants identified visual media as helping them effectively access preventive health information.

As one participant explained:

> *“Maybe something graphic like with the smoking ads when they dissected the lung and you could see the tar. So maybe dissecting a brain and just showing decades of alcohol abuse is going to maybe shrink your brain down to a Mandarin or something.”* – Sydney, Female, 36-45 years

Another participant described using cooking videos to help them eat well:

> *“I watch cooking videos, they’re good, I enjoy those. Get them on. It’s great. Especially if they’ve got a fun personality you’re like, oh yeah, I’ll make it with you.”* – Brisbane, Female, 26-35 years

##### Not boring or scary

Participants explained messages that were “fun” rather than “boring” would be more engaging:

> “*I think something fun and easy to remember. Something catchy and something that I want to listen or I want to watch more about it. So, something not too boring*.” – Brisbane, Male, 36-45 years

From a different angle, another participant described anything too scary as deterring them from giving it their attention:

> “*I think for me, just, like, seeing the word “cancer,” if you just walk by a clinic, it just scares you so much that you don’t want to research into it.*” – Hobart, Male, 26-35 years

#### Sharing preventive health information with others

Many participants offered opinions on the act of sharing preventive health information, positioning themselves not only as recipients of such information, but as active agents in its dissemination with peers and wider community. One participant explained they made a point of sharing information on social media about cancer screening each year to motivate others to undertake screening:

> *“It’s really quite funny actually because I put it on Twitter and I put it on Facebook and I sort of say, “My birthday presents have arrived today. I think it’s important that you test.” Three or four of my friends said, “We’re prompted to test.” The three of them had bowel cancer. I do it every time I get my kit. I do then, I do it when I get the results back and say ‘good news I’m negative’*.” – Melbourne, Male, 56-65 years

However, this proactive approach to information sharing was not universal, with some participants describing a selectivity or reluctance to share. For example, one participant reported only sharing information if someone asked:

> *“Yeah, I guess if they ask the information, but personally I think I wouldn’t be sharing information on these things unless they asked for it*.” – Hobart, Female, 26-35 years

Another participant is careful to only share information they feel confident is legitimate:

> *“So if I’m going to pass some information to the next person, I need to know that this information is authentic and it’s true. So, I go out of my way to make sure that I know that this information is true before I could tell the next person about it.”* – Online FG2, Female, 18-25 years

Interestingly, the varied factors participants identified as important when receiving preventive health information were not necessarily adhered to when sharing preventive health information amongst their networks. One participant, for example, described frequently sharing preventive health information with others, and seeing this as an important thing to do even if the information is not directly relevant to her:

> *“I share all the time. I do. If I hear something that I don’t know, I’m always telling somebody else in case it might be useful, it might not be useful for me, but it might be useful for somebody else. Because I have a lot of contact with people that don’t have a lot of resources, so it’s really important that you share what’s around. …it’s my way of how you get that ball rolling of sharing information.”* – Hobart, Female, 56-65 years

Another participant reported sharing someone else’s information on social media without researching it further, but feeling that as they were sharing a person’s experience it was not as important to verify the information:

> *“I mean, I have posted in the past, I reposted… Someone who had breast cancer and posted something really poignant on their Facebook saying, ‘I didn’t know alcohol was dangerous.’ It’s like, ‘Wow, that’s really moved me and I’m going to repost that.’ I didn’t look it up, I didn’t look at any articles about alcohol, although of course you can. But yeah, I just reposted it straight away. I guess, it’s a case, it’s a person’s own experience.”* – Online FG1, Male, 46-51 years

## Discussion

This first analysis into consumer perspectives about preventive health information and related behaviour change has identified novel findings offering insights for health promotion practitioners and policymakers, especially given the recent release of Australia’s NPHS. Firstly, the study participants reported diverse motivations for engaging in preventive health behaviours, but most commonly described desires to enhance wellbeing and limit the effects of ageing as key motivators. This highlights an interesting interpretation to the health promotion community which has developed campaigns for over forty years often designed to elicit motivation through fear, rather than through desire for positive outcomes, despite ongoing evidence that this approach has serious limitations [18]. However, more recent research suggests that, despite consumer preference to the contrary, fear-based health promotion campaigns – when used alongside humour, testimonials and other tools - may still be effective [19]. The role of these different tools in delivering salutogenic messages focused on wellbeing and ageing well has not received the same level of attention, despite the salutogenesis and the wellbeing economy being clearly aligned with health promotion [14]. Instead, this population interest in longevity and wellbeing may be capitalised upon by for-profit companies for products and services that overlook, dismiss or underplay the benefits of the foundational behavioural changes outlined in the NPHS [20]. With this in mind, health promotion agencies must explore the beliefs that underpin these motivations and identify their own messaging opportunities to leverage them for better preventive health outcomes in the community.

The critical importance of personalised information and applicability to an individual’s circumstances was also clearly outlined in this study. It is well-established that health promotion has a broad remit extending beyond dissemination of health information to include any activity aimed at enabling people to increase control over and to improve their health [21]. In line with this definition, health promotion for preventive health change requires action in policy, structural environments, community education and health services [22]. It is the health services, in particular, that were highlighted by the study participants as lacking. More specifically, access to guidance from qualified health professionals to support each individual to integrate behaviour change within their life and circumstance is required. While Australia has universal health care, increased costs of delivering primary healthcare has not been matched by government funding [23] and leading public health organisations contend there is insufficient protected funding dedicated to the delivery of preventive health [24]. The outcome of this situation, as reflected amongst our study participants, is that individuals have access to information about the importance of changing risky behaviours, but limited support from the appropriate primary care practitioner in enacting that change when they have limited capacity to do so independently. Research has explored the relative cost benefits of digital and other novel preventive health care delivery methods that incorporate tailored support from qualified health professionals while still achieving a broad community reach [25], but further investment in these opportunities – and in preventive health more generally – appears needed for government to fulfil NPHS objectives [24].

Another layer of complexity with regards to supporting preventive health behaviour change in the community is the role of peers as valued and trusted information sources, as reported by study participants. Social networks can provide important channels of communication and information dissemination [26]. Friends, family and work colleagues are trusted health information sources for the general population when an individual’s information needs coincide with that of their peer. However, information sharing in social networks can also be stymied by fear of social stigma and the perceived credibility of their family or friend’s knowledge and beliefs [27, 28]. Yet, in this current age of misinformation and disinformation [12, 13, 29], peer information sources should be leveraged by the health promotion community rather than dismissed. In particular, there is a need to better understand how peer relationships and social networks can be used to propagate accurate information rather than spread false or misleading information. While some participants described feeling confident and motivated to share preventive health information with their peers, others reported being reticent to do so. For this reason, future research should explore the characteristics of health promotion marketing campaigns that improve their shareability and relatability for the community.

### Limitations

This study is limited by recruitment through social media, which creates sampling bias. However, this is offset by the diversity in age, gender, and geography of study participants. In addition, the face-to-face setting and health-related topics of the discussion groups may have elicited social desirability bias amongst participants, who may have held back opinions or experiences for fear of social judgement. Future studies should employ a diversity of mixed methods to address this is unavoidable limitation of face-to-face research.

## Conclusion

Despite a relatively high level of preventive health literacy, Australians are still challenged when trying to change health behaviours important for preventive health. There is a need for better access to qualified experts to assist with adapting and implementing general health information within their unique life circumstances. As such, governments and health promotion agencies should ensure their programs and services deliver such expert advice. There is also a need to develop preventive health content which disseminates preventive health information in a manner accessible and relatable to the community but which also leverages the social networks individuals use to share and better understand health information.

## Funding acknowledgment

This work was supported by an Australian Research Council Future Fellowship awarded to AS (FT220100610). HF’s postdoctoral appointment is also partially funded through this Fellowship.

## Author contributions

Amie Steel: Conceptualisation, Methodology, Data curation, Formal analysis, Data collection, Project administration, Writing – original draft, Writing – review and editing, Resources, Visualisation, Supervision. Hope Foley: Methodology, Formal analysis, Data collection, Project administration, Writing – review and editing. Jon Adams: Methodology, Formal analysis, Data collection, Writing – original draft, Writing – review and editing.

## Statements and Declarations

### Data availability

The data that support the findings of this study are not openly available due to reasons of sensitivity and are available from the corresponding author upon reasonable request.

### Ethics approval and consent

The study was conducted in accordance with the Helsinki Declaration, and ethics approval was granted by the University of Technology Sydney Human Research Ethics Committee (ETH24-9111). Informed consent was granted by participants prior to participation.

