## Supplementary Information File (Figure S1, Table S1, Methods S1) for "Translating motivations, barriers, support and information into health behaviour change: community members’ response to Australia’s National Preventive Health Strategy"

**For manuscript:**

**Correspondence to:**

Prof Amie Steel, amie.steel [at] scu.edu.au

Dr Hope Foley, hope.foley [at] scu.edu.au

**Contents:**

Details of recruitment and final participant selection

Page 2: Supplementary Figure S1. Flow chart of recruitment, expressions of interest (EOIs) and sample selection

Page 3: Supplementary Table S1. Participant characteristics (detailed view)

Focus group materials (Supplementary Methods S1)

Page 4: Discussion guide

Page 6: Infographic

**Figure S1. Flow chart of recruitment, expressions of interest (EOIs) and sample selection**

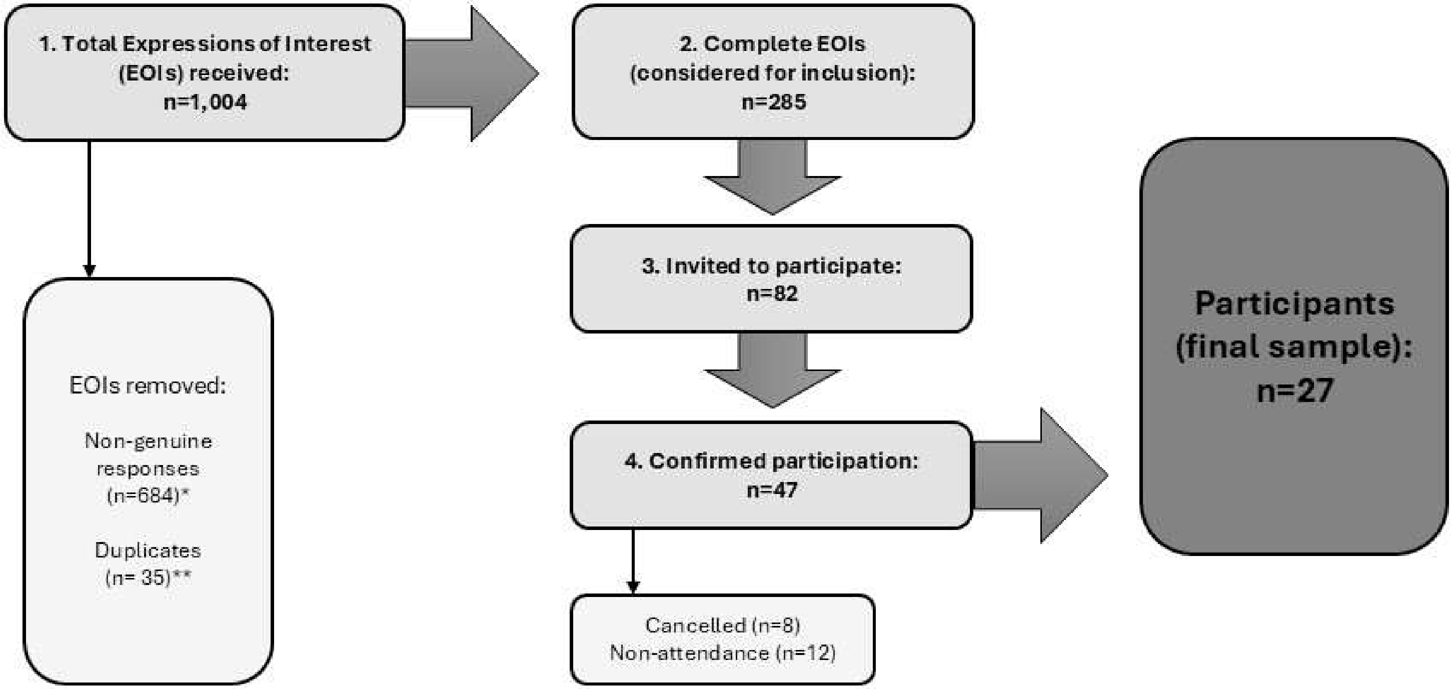


*Responses considered non-genuine were those with international phone numbers, landline phone numbers with area codes not corresponding to the State provided, or those using the same contact details to register for multiple locations.
**Additional responses from potential participants who had submitted multiple identical EOIs.

**Supplementary Table S1. Participant characteristics (detailed view)**

| No. | Gender | Age Range | Use of complementary medicine and health care* | | | | State | Locality | FG Location |
|---|---|---|---|---|---|---|---|---|---|
| | | | Product | Practitioner | Practice | None | | | |
| P1 | Male | 36-45 | - | - | X | - | Qld | Capital city | Brisbane |
| P2 | Non-binary | 26-35 | X | - | - | - | Qld | Capital city | Brisbane |
| P3 | Female | 26-35 | - | - | - | X | Qld | Capital city | Brisbane |
| P4 | Male | 18-25 | - | - | X | - | Qld | Capital city | Brisbane |
| P5 | Male | 36-45 | - | - | - | X | NSW | Capital city | Sydney |
| P6 | Female | 36-45 | X | - | X | - | NSW | Capital city | Sydney |
| P7 | Female | 36-45 | X | X | X | - | NSW | Capital city | Sydney |
| P8 | Female | 46-55 | X | - | X | - | NSW | Capital city | Sydney |
| P9 | Female | 66-75 | - | - | - | X | NSW | Capital city | Sydney |
| P10 | Male | 66-75 | X | - | X | - | NSW | Capital city | Sydney |
| P11 | Female | 56-65 | X | - | - | - | Tas | Capital city | Hobart |
| P12 | Female | 36-45 | - | - | X | - | Tas | Capital city | Hobart |
| P13 | Female | 26-35 | - | - | - | X | Tas | Capital city | Hobart |
| P14 | Male | 26-35 | X | - | - | - | Tas | Capital city | Hobart |
| P15 | Male | 56-65 | - | - | - | X | Vic | Capital city | Melbourne |
| P16 | Female | 18-25 | X | - | X | - | Vic | Capital city | Melbourne |
| P17 | Female | 46-55 | X | - | X | - | Vic | Capital city | Melbourne |
| P18 | Male | 26-35 | X | X | X | - | Vic | Capital city | Melbourne |
| P19 | Female | 56-65 | - | - | - | X | Vic | Capital city | Melbourne |
| P20 | Non-binary | 26-35 | - | X | X | - | Vic | Capital city | Melbourne |
| P21 | Male | 18-25 | X | - | - | - | Vic | Capital city | Melbourne |
| P22 | Female | 26-35 | - | - | X | - | Vic | Capital city | Melbourne |
| P23 | Male | 56-65 | - | X | X | - | Qld | Regional | Online |
| P24 | Male | 46-55 | X | - | X | - | Vic | Regional | Online |
| P25 | Female | 26-35 | X | - | X | - | Vic | Regional | Online |
| P26 | Male | 26-35 | - | - | - | X | Qld | Regional | Online |
| P27 | Female | 18-25 | - | - | X | - | WA | Capital city | Online |

*Use of products (e.g., herbal medicines, supplements), practitioners (e.g., osteopath, acupuncturist or naturopath) and practices (e.g., yoga, meditation) other than mainstream health and medical services; to capture a diverse range of health information experiences.

**Focus Group Discussion Guide for Project:**

### Engagement with publicly-available health information about priority preventive health behaviours: A focus group study amongst the general population [ARC-FF Project Stage E1C, ETH24-9111]

Sample questions for focus group discussions.

*Questions and topics may be edited during the course of the study in response to emergent themes that contribute to addressing the research aims.*

Resources: Table-top graphic icons to use as prompts during discussion, depicting some of the priority health topics (e.g., healthy eating, physical activity, alcohol).

Study Aim: This study explores the perceptions, experiences, values and decision-making that individuals undergo when accessing and using publicly available health information about priority health behaviours outlined in the National Preventive Health Strategy, as shared by CM practitioners through public communication channels and compared to mainstream sources of public health information.

1. Introductory questions

Focus groups will receive an introduction restating the study and participation requirements (as outlined in participant information sheet). The National Preventive Health Strategy will be briefly explained and the priority topics of the Strategy/focus group introduced.

Participants will be asked ice-breaking questions to encourage thinking about the general topic of health promotion, e.g.:

*Can you think of any areas of your life where you try to make healthy choices? In what areas would you like to make healthier choices?*

*What kinds of information have you seen in public places about being healthier?*

2. Accessing health promotion information

Open questions about how health promotion information is generally accessed.

*Where do you go to find information about [preventive health topics]?*

*How do you decide where to go for this information?*

*When do you look for [information source/topic]?*
*[Prompt if needed: Do you search actively for information, or just happen across it?]*

*How do you decide when you need to go elsewhere for such information (e.g., to a health care provider)?*

*Have you encountered any difficulties finding the information you need? What were they?*

*What is important to you when looking for/accessing health promotion information?*

3. <u>Making sense of health promotion information</u>

Open questions about how content, validity, reliability, trustworthiness are assessed.

*How do you assess the information you find to decide whether it is good quality or not?*

*What kind of information do you trust/distrust?*
*(Does it matter who wrote/provided the information? [further questions re: CM practitioners]*
*Does it matter where the information comes from – TV, radio, websites?)*

*Is there anything you do to verify or cross-check the information you come across?*

*If you're not sure about the quality of the information, what do you do next?*

4. <u>Sharing information</u>

*When you find information [online/in public/on TV], do you talk about it with other people?*
*Do you discuss it with your health care providers? [further questions re: CM practitioners]*

*If the information you find about [topic] conflicts with information from your health care provider, what do you do next?*
*Is this different depending on the type of health care provider?*

5. <u>Value of information</u>

*What is it about [participants' preferred sources] that works for you?*

*What do you do with the information you get from [participants' preferred sources/CM practitioner authored sources]?*

*How does [information source] affect your perception/decision-making about [topic/behaviour]?*

*Can you tell us about a time that you found information about being healthy that helped you to make decisions/change your habits? What was it about that information that helped so much?*

6. <u>Information source media</u>

*We recently conducted a survey study which suggests people in Australia are accessing a lot of information about health and being healthy from the television and radio. What are your thoughts on this type of information?*

*How would you compare information from [source type] and [source type]?*
*[further questions re: CM practitioner-authored sources as appropriate]*

**Supporting infographic: Seven preventive health priority areas outlined in the National Preventive Health Strategy, available for viewing by participants throughout discussions.**

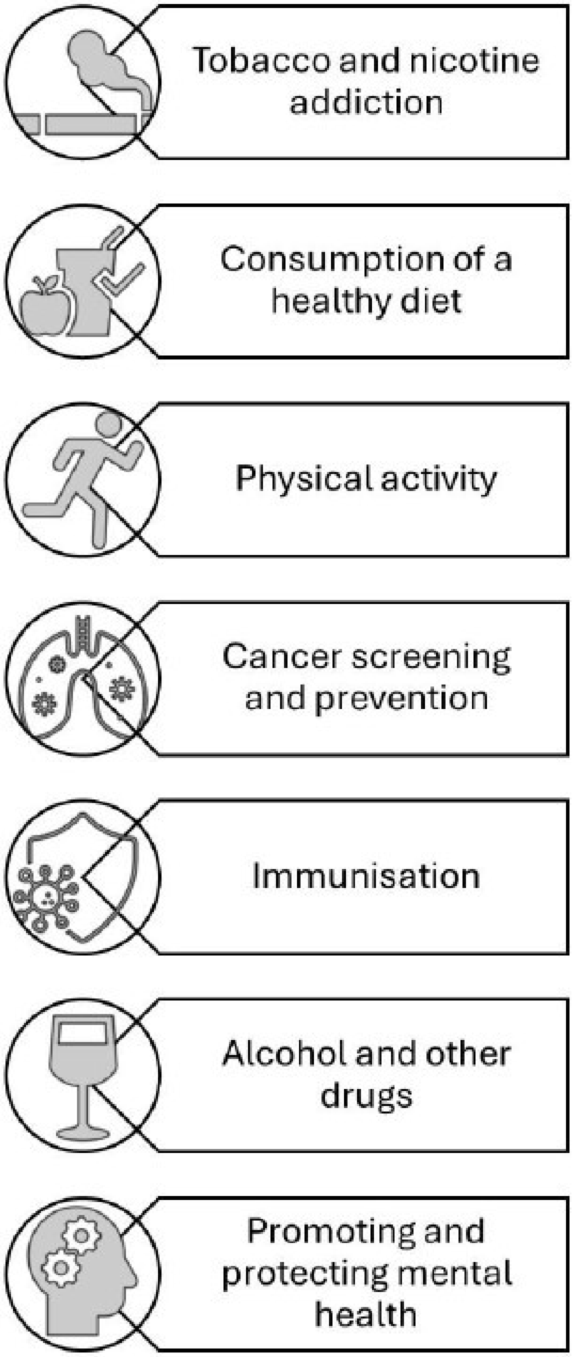
